# The Impact of COVID-19 Testing on College Campuses*

**DOI:** 10.1101/2021.08.16.21262153

**Authors:** Zhuoting Yu, Akane B. Fujimoto, Pinar Keskinocak, Julie L. Swann

## Abstract

**Background:** After moving instruction online for more than a year, many colleges and universities are preparing to reopen and offering fully in-person classes for the Fall 2021 semester. In this paper, we study the impact of weekly testing protocols on college campuses.

**Methods:** An extended susceptible–infectious–removed (SIR) compartmental model was used to simulate COVID-19 spread on a college campus setting. Seven scenarios were evaluated which considered polymerase chain reaction (PCR) and rapid antigen testing kits available at various levels of supply. The infection attack rate (IAR), the number of infections, and the number of tests utilized by the end of the simulation semester are reported and compared.

**Results:** Weekly testing significantly reduces the number of infections compared to when testing is not available. The use of PCR tests results in the lowest infection attack rate and the total number of cases; however, using rapid antigen tests with higher coverage is more effective than using PCR tests with lower coverage.

**Conclusions:** The implementation of COVID-19 testing protocols should be considered and evaluated as using testing allows for identification and isolation of cases which reduces the spread of COVID-19 on college campuses. Even if testing capacity is limited, its partial implementation can be beneficial.

## Introduction

As many colleges are preparing for in-person fall semesters for the 2021-2022 academic year, the safety of the reopening campus remains outstanding. College campuses have been focal points of COVID-19 outbreaks as they house thousands of students that live and interact in indoor spaces for long periods and where social distancing is not often possible, even with restrictions in place (Gressman and Peck [2020], Borowiak et al. [2020]). In this paper, we developed a compartmental model to evaluate the role that periodic weekly testing during the semester has in the spread of COVID on campus. Two types of diagnostic testing, polymerase chain reaction (PCR) and rapid antigen testing kits (Manabe et al. [2020]), are considered in the model. We created various scenarios that use these two different diagnostic testing kits, which are available at various levels of supply.

## Methods

We developed an extension of a susceptible–infectious–removed (SIR) model which contains eleven states. We assumed that the mortality rate for the college student age group is 0, hence there is no dead state in the model. There are three main groups of the states:

1. Susceptible
  i. *S*: susceptible;
2. Infectious
  i. *I*_*a*_: infected and asymptomatic;
  ii. *I*_*s*_: infectious and symptomatic;
  iii. *TFN*_*a*(*s*)_: infected and asymptomatic (symptomatic), has been tested with false negative results;
  iv. 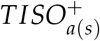 : infected and asymptomatic (symptomatic), has been tested and currently in isolation;
  v. 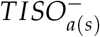: infected and asymptomatic (symptomatic), has been tested and currently not in isolation;
3. Removed
  i. *RC*: Recovered and confirmed, i.e., has been diagnosed previously;
  ii. *RU*: Recovered unknown, i.e., has not been diagnosed previously.

Figure 1 captures the dynamics of the infections in the population. In particular, the following set (Equations 1 - 9) of ordinary differential equations describe the fraction of individuals in each state over time. We assumed that testing occurs weekly every Monday and that results were available immediately.

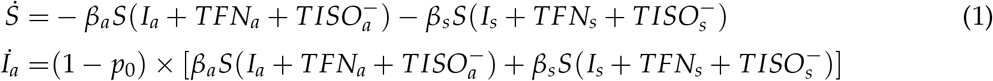

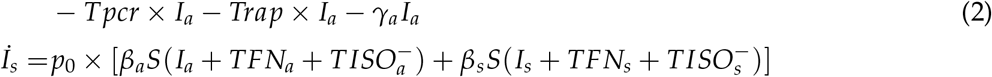

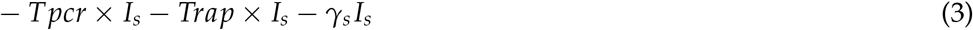

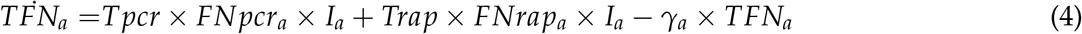

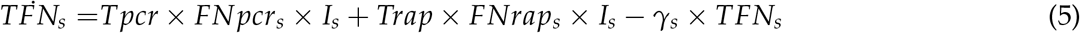

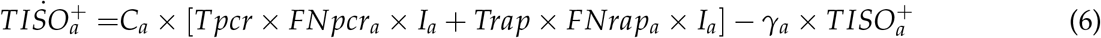

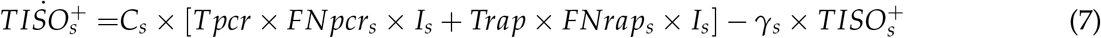

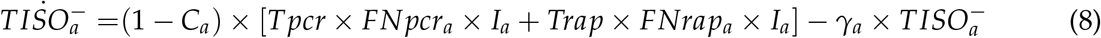

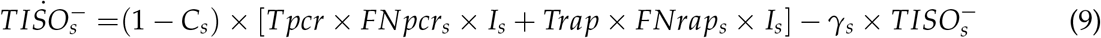

**Figure 1:**
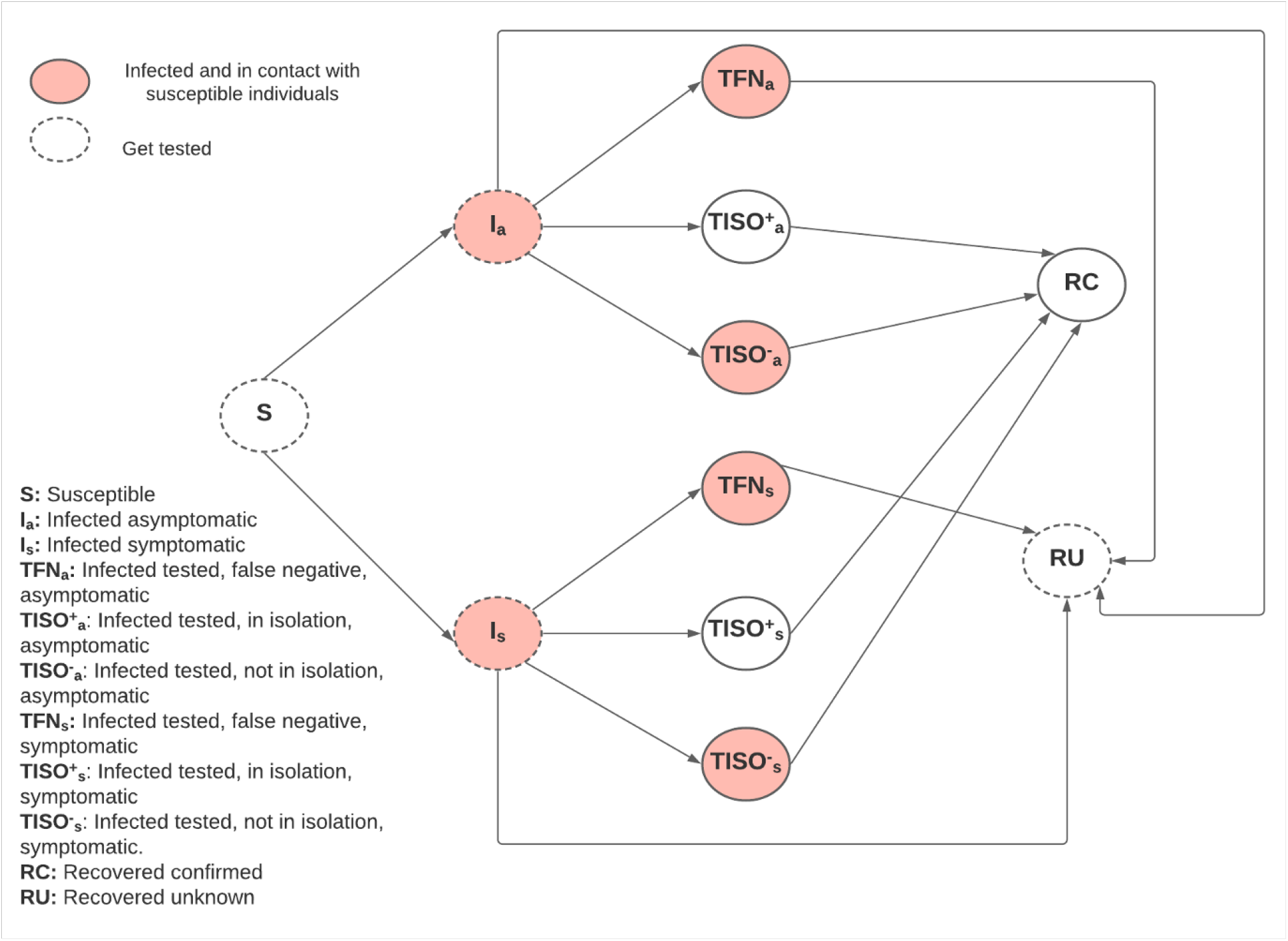
Diagram of the extended SIR model.

Table 1 summarizes the parameters and their values in the model.

**Table 1:** Summary of parameters.

| Parameter | Description | Value |
| --- | --- | --- |
| $\beta_a, \beta_s$ | Transmission rate due to contact between susceptible and asymptomatic/symptomatic infected subjects | 0.1, 0.2 |
| $\gamma$ | Rate of recovery of an infected individual | 0.1 |
| $p_0$ | Proportion of infections that are symptomatic | 0.5 |
| $C_a, C_s$ | Isolation compliance rate for asymptomatic/symptomatic infected and tested individuals | 0.7, 0.9 |
| $FN_{pcr_a}, FN_{pcr_s}$ | False negative rate of a PCR test for asymptomatic/symptomatic patients | 15%, 15% |
| $FN_{rap_a}, FN_{rap_s}$ | False negative rate of a rapid test for asymptomatic/symptomatic patients | 30%, 30% |
| $FP_{pcr_a}, FP_{pcr_s}$ | False positive rate of a PCR test for asymptomatic/symptomatic patients | 5%, 5% |
| $FP_{rap_a}, FP_{rap_s}$ | False positive rate of a rapid test for asymptomatic/symptomatic patients | 10%, 10% |
| $T_{pcr}$ | Percentage of students that have access to PCR tests | Defined in each scenario |
| $Trap_a, Trap_s$ | Percentage of students that have access to rapid antigen tests | Defined in each scenario |

We modeled 7 scenarios that use PCR or rapid antigen testing available for various proportions of the student population per week. PCR tests have higher sensitivity and specificity than rapid antigen tests. Only students who have not been previously diagnosed are eligible for testing.

There is no testing available in the baseline scenario.

- Baseline: No testing
- PCR 100: PCR tests available to 100% of the eligible student body per week.
- RA 100: Rapid antigen tests available to 100% of the eligible student body per week.
- PCR 50: PCR tests available to 50% of the eligible student body per week.
- RA 50: Rapid antigen tests available to 50% of the eligible student body per week.
- PCR 33: PCR tests for 33% available to the eligible student body per week.
- RA 33: Rapid antigen tests available to 33% of the eligible student body per week.

To initialize the model, we assumed that at the time of arrival 89.5% of the student body was susceptible to infection, 0.25% was infected symptomatic, 0.25% was infected asymptomatic, and 10% was recovered unknown. The student body included 15,000 students and the simulation time period considered in the model was from January 18th, 2021 to May 5th, 2021 (Spring 2021). The outcomes of the model include the infection attack rate (IAR), which is equivalent to the percentage of the population that was infected by the end of the simulation, weekly and total number of infections, and total number of tests utilized.

## Results and Discussion

We report the resulting IAR, the number of infections, and the number of tests utilized by the end of the simulation under the various scenarios in Table 2. The results show that having any level of testing significantly reduced the IAR and infections compared to the baseline where testing was not in place (up to a 91% decrease in infections). Using PCR tests resulted in the lowest IAR; however, using rapid antigen tests with higher coverage was better than using PCR tests with lower coverage. Figure 2 shows the IAR progression as the semester advances.

**Table 2:** Summary of the outcomes by scenario.

| Scenario | Cases | IAR (%) | Tests Used |
| --- | --- | --- | --- |
| Baseline | 4,172 | 27.81 | - |
| PCR_100 | 381 | 2.54 | 223,226 |
| RA_100 | 565 | 3.77 | 223,127 |
| PCR_50 | 755 | 5.03 | 111,524 |
| RA_50 | 1,064 | 7.09 | 111,519 |
| PRC_33 | 1,198 | 7.99 | 73,602 |
| RA_33 | 1,556 | 10.37 | 73,627 |

**Figure 2:**
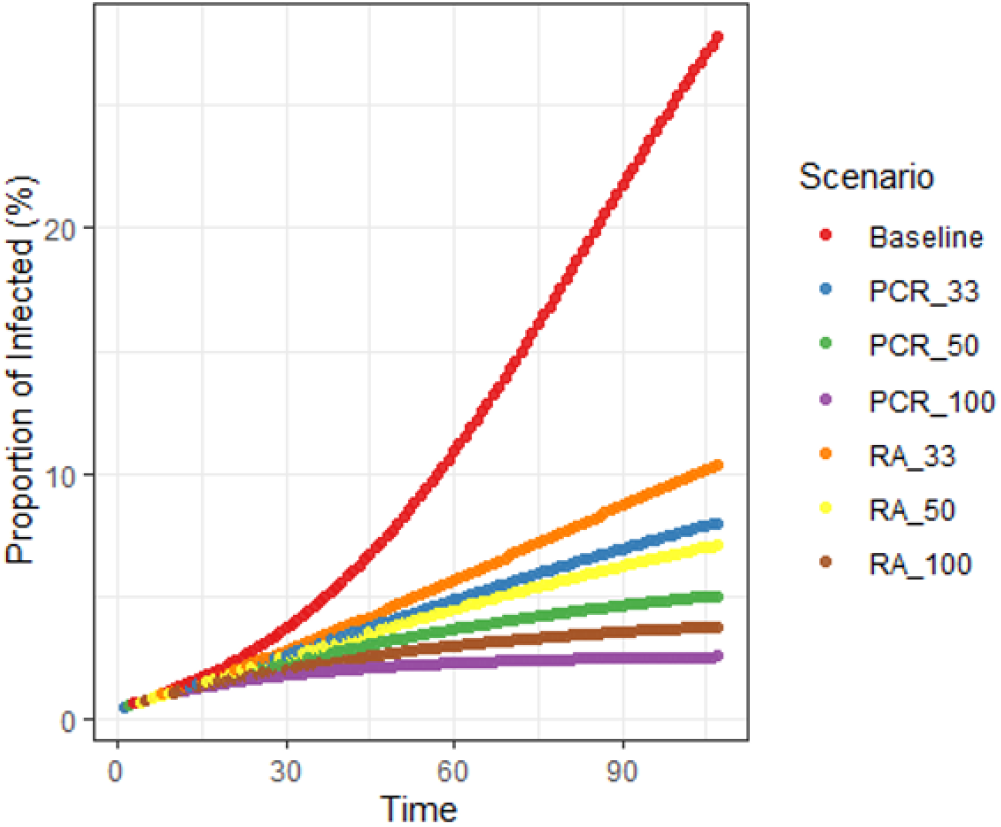
Diagram of the extended SIR model.

The number of new cases every week is displayed in Figure 3. When there was no testing in place (baseline), case incidence rapidly grew week by week. On the other hand, when testing was utilized we see that weekly incidence was controlled and it decreased each week, with the exception of when rapid antigen testing was available to 33% of the student body. In that case, weekly incidence slightly increased but remained controlled.

**Figure 3:**
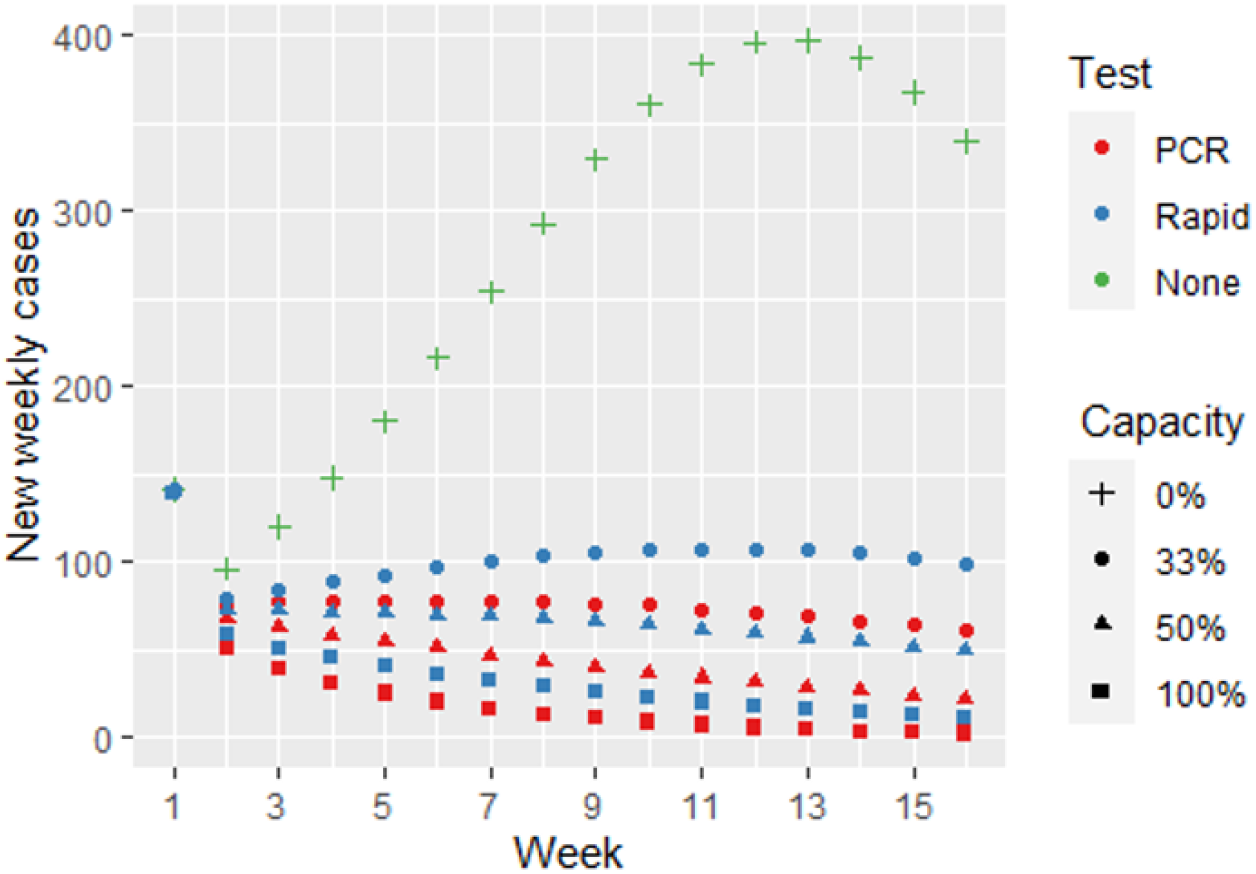
Diagram of the extended SIR model.

The use of testing allows the identification and isolation of active infections. Figure 4 shows the proportion of active cases that were identified and isolated every week via testing. When PCR tests were available to 100% of the student body, the testing protocol was able to identify approximately 70% of the total infections and isolated approximately 55% of the total infections. On the other end, when rapid antigen tests were available to 33% of the student body, approximately 30% of total infections were identified while approximately 25% were isolated.

**Figure 4:**
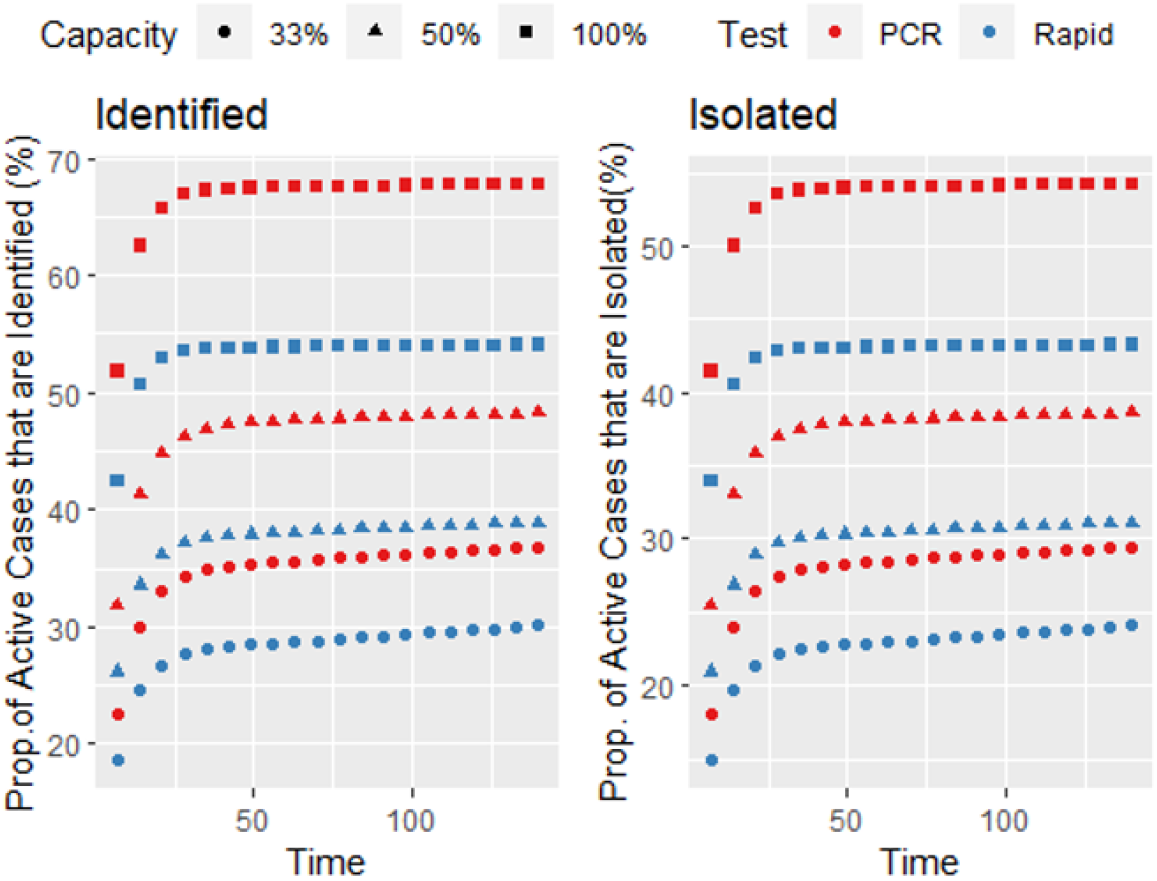
Diagram of the extended SIR model.

## Conclusion

The use of testing (PCR or rapid antigen) in college campuses to identify and isolate active COVID cases significantly reduces the number of infections and helps controlling outbreaks on campus. The implementation of COVID-19 testing protocols, even if testing capacity is limited, should be considered and evaluated as using tests of less accuracy and in lower quantities is still beneficial.

## Data Availability

The manuscript uses publicly available data.

